# Sotatercept is Associated with Improved Lung Function in Sarcoidosis-Associated Pulmonary Hypertension

**DOI:** 10.1101/2025.10.03.25337126

**Authors:** Hooman D. Poor, Elliot Eisenberg, Shyla Saini, Simone Hannah-Clark, Jimmy Zhang, Alison G. Lee, Gregory Serrao, Charles Powell, Corey E. Ventetuolo, Maria Padilla

## Abstract

Sarcoidosis is often complicated by the development of pulmonary hypertension (PH). Sotatercept, a ligand trap for selected transforming growth factor β superfamily members, is a novel, highly effective therapy approved for the treatment of pulmonary arterial hypertension. We conducted a retrospective study of patients with biopsy-proven sarcoidosis who received sotatercept for the treatment of sarcoidosis-associated pulmonary hypertension (PH) to assess the effect of sotatercept on lung function. Nine patients were included in the study. Median age was 64 years (IQR 56 to 71), seven patients (78%) were female. All patients were receiving chronic background immunosuppression, and eight patients (89%) were receiving background pulmonary vasodilators. Pulmonary function testing following initiation of sotatercept (median 4.4 months, IQR 2.9 to 8.0 months) demonstrated improvement in lung function in all patients: FEV1 increased 150 mL (13%, IQR 65 to 340 mL), FVC increased 170 mL (11%, IQR 65 to 285 mL), DLCO increased 0.95 mL/min/mmHg (14%, IQR 0.30 to 2.59 mL/min/mmHg), and FEF25-75% increased 0.18 L/s (55%, IQR −0.04 to 0.75 L/s). One patient had near complete radiographic resolution of pulmonary infiltrates. Seven patients (78%) had stable or decreasing background immunosuppression. In patients with sarcoidosis-associated PH, sotatercept was associated with improvement in lung function, and in particular, markers of small airways disease.

---

Sarcoidosis, a multisystemic disease characterized by noncaseating granulomatous inflammation, is often complicated by the development of pulmonary hypertension (PH) [1]. Sotatercept, a ligand trap for selected transforming growth factor β (TGF-β) superfamily members, is a novel, highly effective therapy approved for the treatment of pulmonary arterial hypertension (PAH) [2]. Here, we report a study of patients treated off-label with sotatercept for precapillary PH associated with sarcoidosis and their unexpected improvement in lung function. The Mount Sinai Institutional Review Board gave ethical approval for this work (IRB approval 19-00263); all patients were clinically counseled about the off-label use of sotatercept.

The first patient, which informed the use of sotatercept in subsequent cases, was a woman in her 40’s with sarcoidosis-associated PH (mean pulmonary artery pressure [mPAP] 48 mmHg, pulmonary vascular resistance [PVR] 7.6 WU, cardiac index [CI] 2.8 L/min/m^2^) and moderate right ventricular (RV) dysfunction, who exhibited functional class (FC) III symptoms despite triple oral PAH therapy (tadalafil, macitentan, selexipag). Oxygen saturation breathing room air was 92% with exertional desaturation to 84%. She was started on azathioprine 100 mg daily due to worsening lung function (FVC 57% predicted, FEV1 48% predicted, and DLCO 50% predicted) and progressive nodular consolidations on chest CT (Figure 1). Given her prior experience with steroids, the patient did not wish to initiate prednisone. An indwelling pleural catheter (IPC) was placed to drain reaccumulated transudative pleural effusions (Figure 1A; filled triangle). She drained approximately one liter of pleural fluid twice a week, the amount unchanged after an increase in diuretics. Repeat pulmonary function tests (PFT) after three months of azathioprine (Figure 1A) and repeat chest CT after six months of azathioprine (Figure 1B) were unchanged. Given her poor exercise capacity, persistent RV dysfunction, and precapillary PH (mPAP 45 mmHg, PVR 4.8 WU, CI 3.3 L/min/m^2^), sotatercept was initiated. Within two weeks, the patient noted marked improvement in exercise capacity (FC II). Her IPC stopped draining and was removed one month after sotatercept initiation (Figure 1A, hollow triangle). Fourteen months after starting sotatercept, her PFTs (Figure 1A) normalized with FVC 106% predicted, FEV1 98% predicted, and DLCO improved to 70% predicted. Repeat chest CT revealed significant decrease in perilymphatic nodules and areas of consolidation, with near complete resolution of the left pleural effusion (Figure 1C). Gas exchange also improved (98% breathing room air, 94% with exertion). Repeat hemodynamics demonstrated decreased mPAP but essentially unchanged PVR (mPAP 31 mmHg, PVR 5.0 WU, CI 2.2 L/min/m^2^).

**Figure 1:**
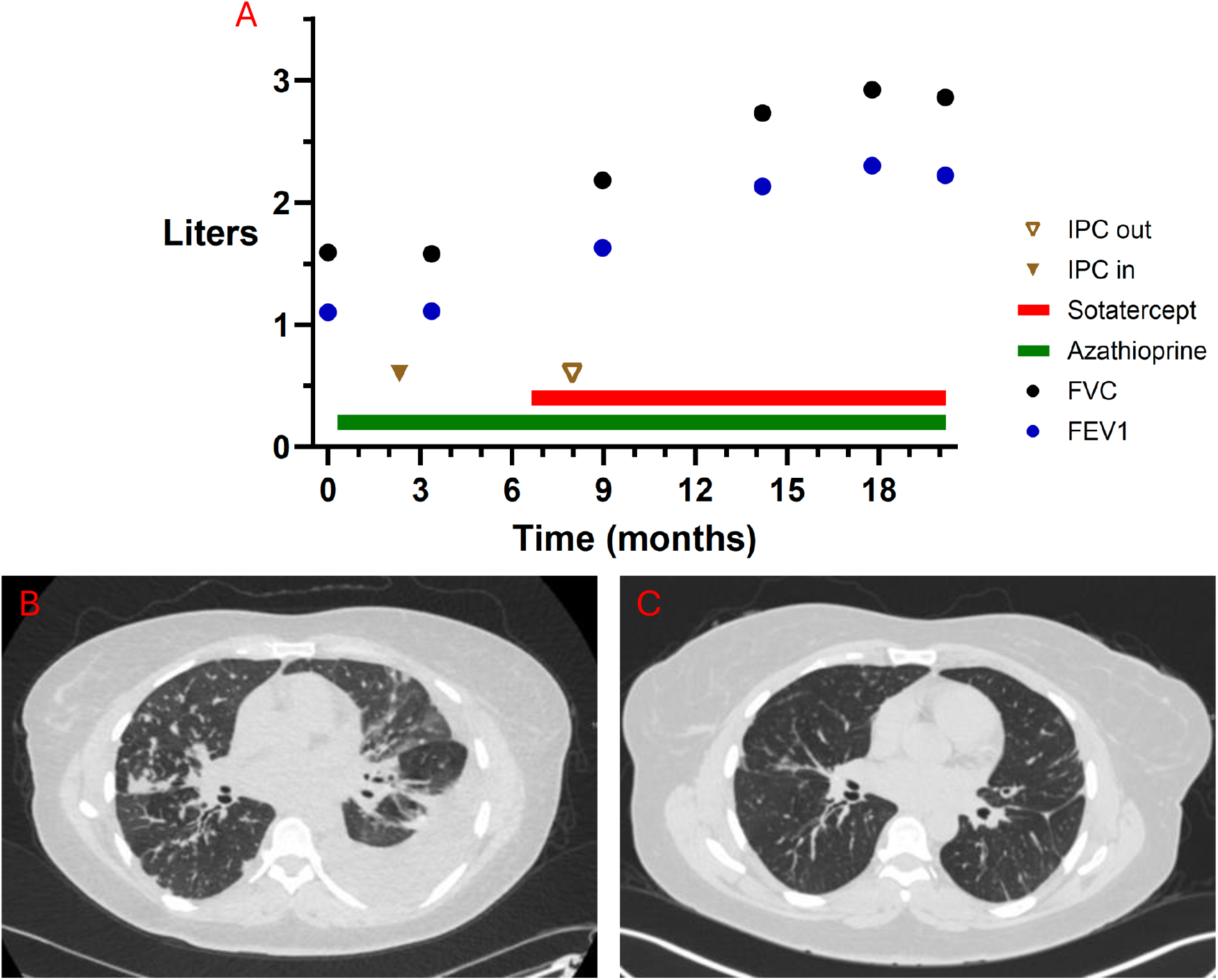
Spirometry and imaging in Patient 1. **A. Spirometry trend over time**. There was no change in spirometry after three months of treatment with azathioprine alone, but progressive improvement in spirometry soon after initiation of sotatercept. Filled triangle represents placement of indwelling pleural catheter (IPC) and hollow triangle represents removal of IPC. **B and C. Noncontrast chest computed tomography (CT) images before and after initiation of sotatercept**. *(B)* CT scan after six months of therapy with azathioprine demonstrating diffuse subpleural and peribronchovascular nodules bilaterally and a moderate left pleural effusion. These findings were unchanged compared to the CT scan done six months prior, just before initiation of azathioprine (not shown). *(C)* CT scan after 11 months of sotatercept demonstrating marked reduction in nodules and near complete resolution of the left pleural effusion. CT images at the level of the right secondary carina.

We went on to treat eight additional patients with biopsy-proven sarcoidosis and precapillary PH demonstrated on right heart catheterization at a single center. The median time since sarcoidosis diagnosis was 23 years (interquartile range [IQR] 17 to 29). Seven patients (78%) were female and median age was 64 years (IQR 56 to 71). One patient (11%) was on background PAH monotherapy, four (44%) on dual therapy, and three (33%) on triple therapy; one patient (11%) could not tolerate pulmonary vasodilators due to systemic hypotension. All patients were receiving chronic immunosuppression. During the six months prior to sotatercept initiation until the last PFT measurement, immunosuppression intensity was increased for two patients (22%), decreased for four patients (44%), and remained the same for three patients (33%).

Lung function in the year prior to sotatercept was worsening for all patients (Figure 2). PFTs following initiation of sotatercept (median 4.4 months, IQR 2.9 to 8.0) demonstrated improvement in lung function in all patients. Figure 2 demonstrates increases in FEV1 (150 mL [13%], IQR 65 to 340), FVC (170 mL [11%], IQR 65 to 285), DLCO (0.95 mL/min/mmHg [14%], IQR 0.30 to 2.59), and FEF25-75% (0.18 L/s [55%], IQR −0.04 to 0.75) after sotatercept initiation. There was no overall change in FEV1/FVC; however, all three patients with decline in FEV1/FVC had significant increases in FVC (120, 230, and 170 mL), and FEV1 (20, 20, and 150 mL), suggesting that despite a decline in FEV1/FVC, airway function improved and distal airway closure was delayed. Notably, those three patients were the only to have decreases in FEF25-75% after sotatercept initiation, also consistent with delay in distal airway closure. For six patients (66%), FEV1 after sotatercept initiation was the highest measured over the prior four years.

**Figure 2:**
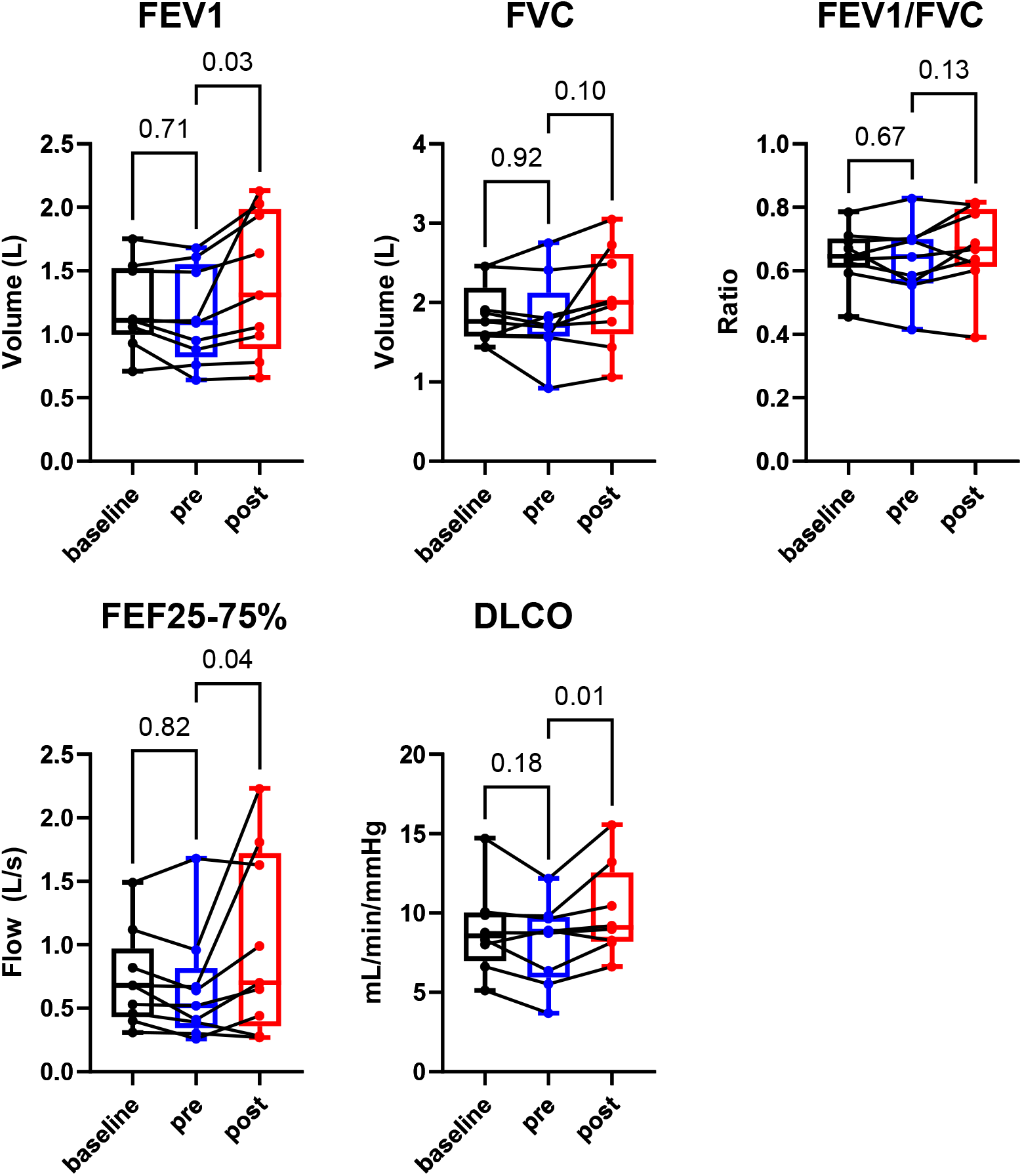
Change in pulmonary function testing (PFT) over time, at baseline (within one year), before, and after initiation of sotatercept. There are trends of worsening lung function between baseline and pre-sotatercept PFTs (median time between PFTs 4.2 months, IQR 3.6 to 5.0). Compared to pre-sotatercept PFTs, post-sotatercept PFTs demonstrate increases in FEV1, FEF25-75%, and DLCO, with trend towards improvement in FVC and FEV1/FVC. Median time between pre-sotatercept and post-sotatercept was 8.9 months (IQR 4.3 to 12.6).

While the intention of treatment with sotatercept in these patients was to improve the pulmonary vascular component of sarcoidosis, we noted improvement in small airways disease and parenchymal lung disease. Although it is possible that the patients’ improvement in lung function could have occurred spontaneously or with changes in immunosuppression, we note that seven patients demonstrated improvement despite stable or even decreasing background immunosuppression. Additionally, many of these patients had evidence of deteriorating lung function preceding sotatercept administration.

We hypothesize that these findings suggest that sotatercept has anti-inflammatory effects on the granulomatous inflammation of sarcoidosis, especially in the small airways. Micro-CT scanning and histology support granulomatous remodeling as a cause of small airways loss in fibrotic sarcoidosis [3]. Sotatercept’s efficacy in PAH is thought to result primarily from its inhibition of activin signaling, leading to decreased cell proliferation and differentiation in the pulmonary vasculature [4]. Beyond vascular remodeling, ligands of the TGF-β superfamily are key regulators of inflammation. Activins drive macrophage activation, cytokine production (including IL-1β, IL-6, and TNF-α), and tissue infiltration [5]. Granulomatous inflammation via activated epithelioid macrophages is a hallmark of sarcoidosis, a multisystem disorder commonly with pulmonary involvement and occasionally pleural involvement [6]. Activin A levels are elevated in patients with sarcoidosis, and the TGF-β/Smad pathway has been implicated in pulmonary sarcoidosis [7-9]. In animal models of PAH, a sotatercept analogue reversed proinflammatory gene expression in macrophages and normalized pulmonary macrophage infiltration [10].

Limitations of this report include the small sample size, lack of a control group, and lack of formal exercise testing, which may have provided information about dynamic cardiopulmonary interactions in these patients with sotatercept treatment. In summary, we present a study of sarcoidosis patients with PH who demonstrated small airways and parenchymal improvements following initiation of sotatercept. Sotatercept could be a potential treatment for pulmonary sarcoidosis, with or without PH, and further clinical research is warranted.

## Data Availability

All data produced in the present study are available upon reasonable request to the authors

## Notes

**Sources of support:** C.E.V. R01HL174007; U54GM115677-09S3 (PD: Rounds); AHA 24IPA1275127

**Conflicts of Interest:** H.D.P. advisory board for Merck E.E. none S.S. none S.H-C. none J.Z. none A.G.L. none G.S. none C.P. none C.E.V. consulting fees from Merck and Regeneron Pharmaceuticals; advisory boards for Janssen Pharmaceuticals and Merck; institution receives clinical trial support from Pulmovant, Gossamer Bio, Pfizer, Tenax Therapeutics, and Merck M.P. consulting fees from Boehringer Ingelheim and United Therapeutics

### Competing Interest Statement

Conflicts of Interest:
H.D.P. advisory board for Merck
E.E. none
S.S. none
S.H-C. none
J.Z. none
A.G.L. none
G.S. none
C.P. none
C.E.V. consulting fees from Merck and Regeneron Pharmaceuticals; advisory boards for Janssen Pharmaceuticals and Merck; institution receives clinical trial support from Pulmovant, Gossamer Bio, Pfizer, Tenax Therapeutics, and Merck
M.P. consulting fees from Boehringer Ingelheim and United Therapeutics

### Funding Statement

C.E.V. has received funding from: R01HL174007; U54GM115677-09S3 (PD: Rounds); AHA 24IPA1275127

### Author Declarations

The Mount Sinai Institutional Review Board gave ethical approval for this work (IRB approval 19-00263)

